# Comparative effects of viral transport medium heat inactivation upon downstream SARS-CoV-2 detection in patient samples

**DOI:** 10.1101/2020.07.30.20164988

**Authors:** Jamie L Thompson, Angela Downie Ruiz Velasco, Alice Cardall, Rebecca Tarbox, Jaineeta Richardson, Gemma Clarke, Michelle Lister, Hannah C Howson-Wells, Vicki M Fleming, Manjinder Khakh, Tim Sloan, Nichola Duckworth, Sarah Walsh, Chris Denning, C. Patrick McClure, Andrew V Benest, Claire H Seedhouse

## Abstract

The COVID-19 pandemic, which began in 2020 is testing economic resilience and surge capacity of healthcare providers worldwide. At time of writing, positive detection of the SARS-CoV-2 virus remains the only method for diagnosing COVID-19 infection. Rapid upscaling of national SARS-CoV-2 genome testing presented challenges: 1) Unpredictable supply chains of reagents and kits for virus inactivation, RNA extraction and PCR-detection of viral genomes 2) Rapid time to result of <24 hours is required in order to facilitate timely infection control measures. We evaluated whether alternative commercially available kits provided sensitivity and accuracy of SARS-CoV-2 genome detection comparable to those used by regional National Healthcare Services (NHS), and asked if detection was altered by heat inactivation, an approach for rapid one-step viral inactivation and RNA extraction without chemicals or kits. Using purified RNA, we found the CerTest VIASURE kit to be comparable to Altona RealStar system currently in use, and further showed that both diagnostic kits performed similarly in the BioRad CFX96 and Roche LightCycler 480 II machines. Additionally, both kits were comparable to a third alternative using a combination of Quantabio qScript 1-step qRT-PCR mix and CDC-accredited N1 and N2 primer/probes when looking specifically at borderline samples. Importantly, when using the kits in an extraction-free protocol, following heat inactivation, we saw differing results, with the combined Quantabio-CDC assay showing superior accuracy and sensitivity. In particular, detection using the CDC N2 probe following the extraction-free protocol was highly correlated to results generated with the same probe following RNA extraction and reported clinically (n=127; R_2_=0.9259). Our results demonstrate that sample treatment can greatly affect the downstream performance of SARS-CoV-2 diagnostic kits, with varying impact depending on the kit. We also showed that one-step heat inactivation methods could reduce time from swab receipt to outcome of test result. Combined, these findings present alternatives to the protocols in use and can serve to alleviate any arising supply chain issues at different points in the workflow, whilst accelerating testing, and reducing cost and environmental impact.

## Introduction

The current COVID-19 pandemic developed from an outbreak of SARS-CoV-2 virus initially identified in Wuhan, China. Widespread lockdowns and societal behaviour changes were introduced, but even with these in place the economic resilience and surge capacity of healthcare providers worldwide was significantly tested. The UK responded slowly, with established SARS-CoV-2 testing of NHS staff, patients, healthcare providers and other frontline key worker falling behind other comparable economies in Western Europe (1).

Positive detection of the virus, irrespective of a diverse range of symptoms, remains the only method for diagnosing COVID-19 infection with, at the point of writing, antibody detection of uncertain clinical significance (2). Nasopharyngeal or throat swabs can be taken, transported in Viral Transport Medium (VTM) before being treated with heat (to both thermally sterilise the swabbed sample, but additionally to contribute to lysis) or chemical treatment to lyse samples and extract RNA (using reagents such as guanidine thiocyanate, GT). GT has variable reports of effectiveness (3), and the rapid upscale of national COVID-19 testing programmes led to shortages and an unpredictable supply. Isolation of viral RNA using commercially available kits, suited for automation for high throughput systems, had similar supply chain issues and was equally impacted by the COVID-19 pandemic. Either of these instances were equally compounded by fluctuations in commercially available, Public Health England (PHE)-authorised SARS-CoV-2 RT-qPCR tests. For example, the frequently used laboratory RNA extraction kit from QIAGEN became unavailable. Routine diagnostic RNA extraction kits (eg QIAGEN viral RNA extraction kit), normally readily available for NHS laboratories were rationalised and supplies significantly reduced during the pandemic. This prompted significant challenges for laboratories, and although extraction-free (direct PCR) methods are well described (4), side-by-side comparisons between the most available kits and thermocycling machines available to the NHS have not been widely reported.

Viral RNA detection can be used in both community and hospital settings, screening symptomatic, asymptomatic and healthcare workers alike with positive results triggering both clinical and economical decision making. There are multiple nucleic acid extraction and reverse transcriptase PCR (RT-qPCR) methods that can be used to accurately detect viral RNA. Therefore, correct and reliable COVID-19 detection is essential, but it is not clear to what extent changing swab sample treatment will alter downstream detection kit sensitivity and accuracy. RNA extraction methods have resulted in bottlenecks in China, where the significant increase in sample processing time, and failings in constant reagent supply will reduce the ability to perform the high number of tests required to manage pandemics (5). Similarly, although heat inactivation of clinical samples is known to be effective at thermosterilising swab samples (6, 7), it is unclear how this alternative sample processing impacts upon commercially available SARS-CoV-2 RT-qPCR tests.

In order to evaluate local capacity to respond to changes in supply chain of in the availability of SARS-CoV-2 tests, we made side-by-side comparisons of different commercially available SARS-CoV-2 RT-qPCR tests and assessed the influence of extraction-free heat treatment upon downstream detection applications.

## Methods

### Samples

All samples were obtained from nasal or throat swabs in viral transport medium (VTM) from individuals with suspected COVID-19. Samples were anonymised, and surplus clinical material was provided by Nottingham University Hospitals (NUH) NHS trust and Path Links Pathology (PLP). The study was locally assessed to be service development and thus exempt from requirements of additional ethical approval. RNA samples utilised (n=66) had undergone diagnostic testing via RealStar SARS-CoV-2 RT-qPCR kit (Altona Diagnostics). In addition, surplus heat-inactivated VTM samples (n=142) were assessed in extraction-free RT-qPCR assays; aliquots of these samples previously had RNA extracted for clinical diagnostic testing (RealStar® (Altona Diagnostics) or M2000 (Abbott) RealTime SARS-CoV-2 assay testing systems. These samples were heat inactivated at 90 °C – 95 °C for 10 minutes; importantly the total incubation time was 17 minutes to allow for the 7 minute lag time taken for the samples to reach temperature, as tested in our hot block (data not shown). The high temperature was chosen to ensure total inactivation of SARS-CoV-2 and other potential respiratory pathogens (6). RNA was stored at −80 °C and heat-inactivated samples stored at 4 °C short term and −80 °C long term (initial assays demonstrated no change in viral detection following −80 °C storage). SARS-CoV-2 positive, negative and equivocal samples (which we subsequently determined as positive by RNA sequencing methods) were used to compare the RNA extraction and RNA extraction-free methods using various RT-qPCR kits. RT-qPCR cycle numbers generated during this work were compared to those generated diagnostically by the RealStar (NUH) or the Abbott systems (ULH).

### RT-qPCR

The VIASURE SARS-CoV-2 RT-qPCR kit (CerTest Biotech) was used according to manufacturer’s instructions. Each reaction was performed in multiplex for ORF1ab (FAM) and N (ROX) genes with an internal control (HEX) using 15 µl of reaction mix and 5 µl of sample and were run using a LightCycler 480 II (Roche Life Science) or CFX 96 (BioRad) qPCR machine for 45 amplification cycles. Samples were denoted positive where ORF1ab alone or in combination with N showed amplification at <38 cycles. Where N alone was amplified, samples were considered inconclusive. Negative samples showed amplification in internal control sample at <38 cycles only, or in combination with ORF1ab and/or N at >38 cycles.

The Centre for Disease Control and Prevention (CDC, USA) 2019-Novel Coronavirus (2019-nCoV) Real-Time RT-PCR Diagnostic Panel was adopted using either qScript XLT One-Step RT-qPCR ToughMix or UltraPlex 1-Step ToughMix (4x) (Quantabio). Each reaction was performed in single for N1 (FAM), N2 (FAM) genes or RNaseP (FAM) extraction control using 15 µl of reaction mix and 5 µl of sample and were run using a CFX 96 qPCR machine for 45 replication cycles. Samples were denoted positive where both N genes showed amplification at <40 cycles. Where only one N gene amplified, samples were deemed inconclusive. Negative samples showed amplification in RNaseP at <40 cycles only and/or N gene(s) at >40 cycles.

### Data Analysis

C_T_ (cycle threshold) values were determined for each of the genes tested and these were compared with the diagnostic C_T_ values provided by the hospital trusts. All data analysis and graph production were performed using GraphPad Prism 8.3. Where comparisons were drawn between the sensitivity of testing methods ΔC_T_ values for each sample were calculated relative to RealStar S C_T_ value and statistical significance was assessed using a paired t-test. To compare different qPCR machines, median and interquartile range were calculated for C_T_ values obtained. Comparisons were made with clinical diagnostic data to assess the performance of testing methods. Correlations were tested by Pearson’s correlation coefficient and R^2^ values reported.

## Results

### Comparison of kits and qPCR machines

We sought to pre-empt potential problems in local SARS-CoV-2 testing, due to GT and commercial RNA extraction kit supply chain issues by evaluating potential alternatives to the existing standards. We first addressed the need for alternatives to the currently used local diagnostic assay, the Altona diagnostics RealStar SARS-CoV-2 kit, a dual target assay detecting lineage B-βCoV-specific (E) and SARS-CoV-2-specific RNA (S), as well as an internal control (IC). Here, we evaluated the CerTest VIASURE SARS-CoV-2 Real Time PCR Detection kit (herein termed VIASURE), as a potential alternative. In order to do this, we compared blinded RNA samples from patients with a positive (n=26; range of C_T_ values S gene, 12.37-31.22; E gene, 13.26-31.22) or negative (n=9) clinical diagnosis of COVID-19 by the NUH RealStar test with results generated by the VIASURE kit (Table 1, Fig 1A-B).

**Table 1:** Comparison of VIASURE assay using RNA samples run on Bio-Rad CFX96 and Roche Lightcycler 480 II qPCR machines, to clinical RealStar assay

| Probe | Machine | Results (n) |  |  |  | Concordance (%) |
| --- | --- | --- | --- | --- | --- | --- |
|  |  | TP | TN | FP | FN |  |
| ORF1ab | CFX | 26 | 9 | 0 | 0 | 100 |
|  | LC480 | 26 | 9 | 0 | 0 | 100 |
| N | CFX | 26 | 9 | 0 | 0 | 100 |
|  | LC480 | 26 | 9 | 0 | 0 | 100 |

TP, true positive; TN, true negative; FP, false positive; FN, false negative; CFX, Bio-Rad CFX 96; LC480, Roche Lightcycler 480 II

**Figure 1.**
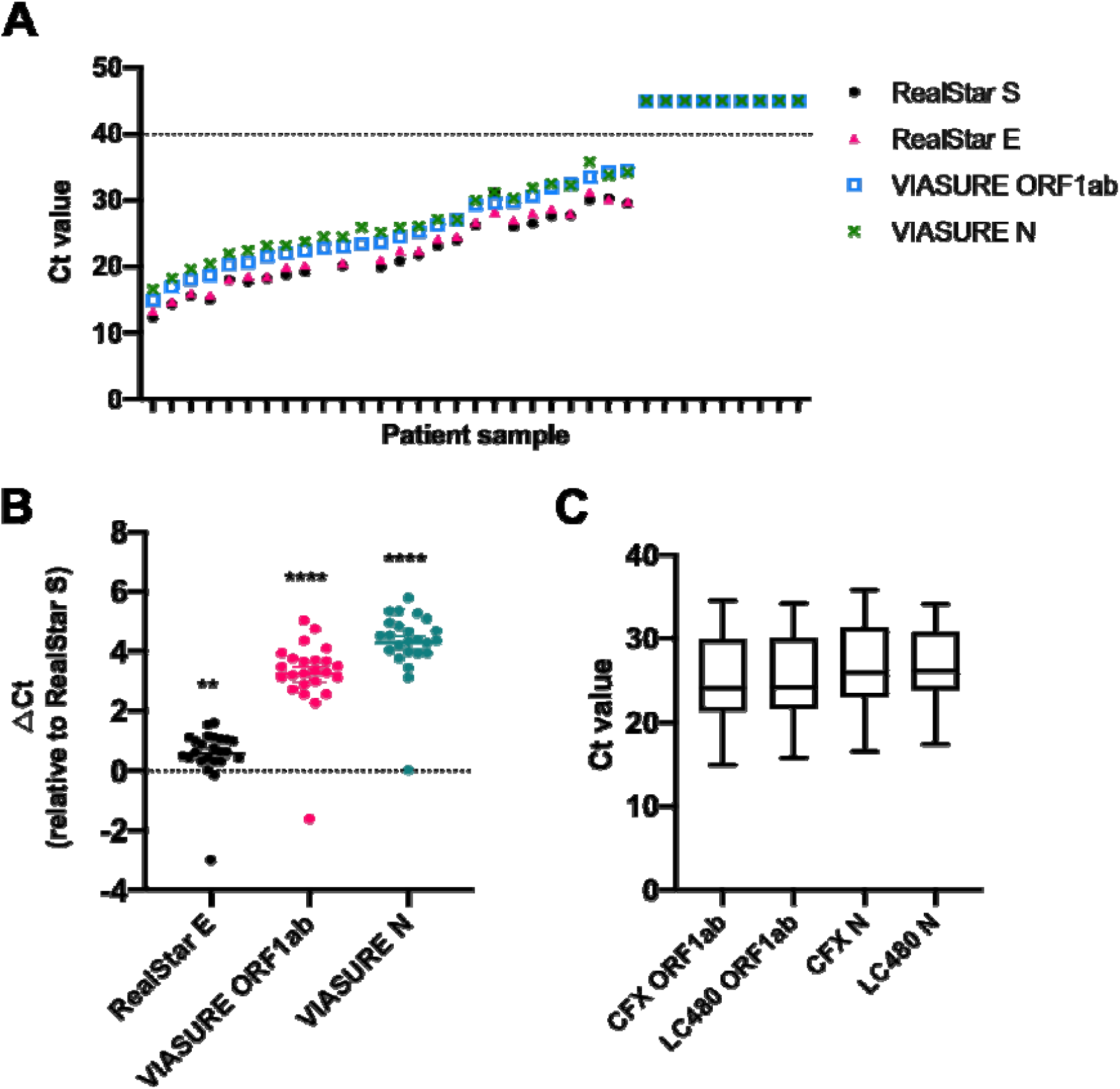
Comparison of VIASURE and RealStar kits using RNA. **(A)** C_T_ values generated from VIASURE ORF1ab and N gene probes and RealStar S and E probes. Individual points represent a single detected SARS-CoV-2 sample. Samples with C_T_ values above the dotted line are interpreted as negative **(B)** ⍰C_T_ for each sample was calculated relative to RealStar S C_T_ value to demonstrate differential probe sensitivity. Statistical differences for each gene were calculated using a paired t-test relative to clinical S (** *p ≤ 0.01, **** p ≤ 0.0001*). **(C)** Comparison of median and interquartile range for C_T_ values obtained using the VIASURE assay on two RT-qPCR machines, Bio-Rad CFX and Roche LC480 with no significant differences found between comparisons. (n=26).

The VIASURE kit uses probes that detect SARS-CoV-2 ORF1ab and N genes as well as an internal control that ensures there is no inhibition of the reaction. Importantly, all samples tested showed perfect concordance with the clinical results (Table 1). Additionally, we were interested in comparing the C_T_ values for both kits as a simplified measure of sensitivity. C_T_ values of ORF1ab and N genes determined by the VIASURE kit as well as RealStar S and E genes were plotted for individual samples (Figure 1A). Overall, RealStar S and E genes showed C_T_ values approximately 3-4 cycles lower than the ORF1ab and N genes, which was apparent in almost all samples. To better illustrate this, C_T_ values for RealStar E, ORF1ab and N genes were normalized to their respective RealStar S value (Figure 1B). The RealStar S gene was significantly more sensitive compared to the RealStar E, ORF1ab and N gene, with a mean C_T_ difference of 0.58, 3.24 and 4.29, respectively.

Next, we asked whether a significant difference in sensitivity of the kit would be observed if a different thermocycler was used. We compared the Bio-Rad CFX96 model currently used by the local clinical laboratory, to the Roche LightCycler 480 II, and saw no difference in average C_T_ value (Figure 1C). Thus, accuracy of VIASURE kit was agnostic of qPCR machines tested and matched the RealStar assay, although the sensitivity was approximately 5- to 10-fold lower.

### Detection of equivocal samples

Having shown perfect concordance between the VIASURE kit and clinical RealStar results using true positive and negative RNA samples, we looked at the performance of the VIASURE kit on borderline (equivocal) samples. We analysed RNA from 18 patient samples reported as inconclusive after analysis with the RealStar system (Table 2, Figure 2). As an additional comparator system, we developed a testing strategy that combined the Quantabio qScript XLT 1-step RT-qPCR mix with CDC-accredited primer/probes specific for the N1 and N2 viral sequences and human RNaseP (RP) sequence, to provide an alternative workflow to commercial SARS-CoV-2 detection kits with combined probe/reaction mixes.

**Table 2:** Sensitivity of RealStar, VIASURE and XLT assays for detection of borderline SARS-CoV-2 samples (n= 18).

| Assay/probe set | Samples detected (n=18) | Average Ct of detected samples |
| --- | --- | --- |
| RealStar S | 0 (0.0 %) | N/A |
| RealStar E | 18 (100.0 %) | 33.3 |
| VIASURE ORF1ab | 15 (83.3 %) | 34.4 |
| VIASURE N | 15 (83.3 %) | 33.9 |
| Quantabio XLT N1 | 13 (72.2 %) | 34.2 |
| Quantabio XLT N2 | 15 (83.3 %) | 33.8 |

**Figure 2.**
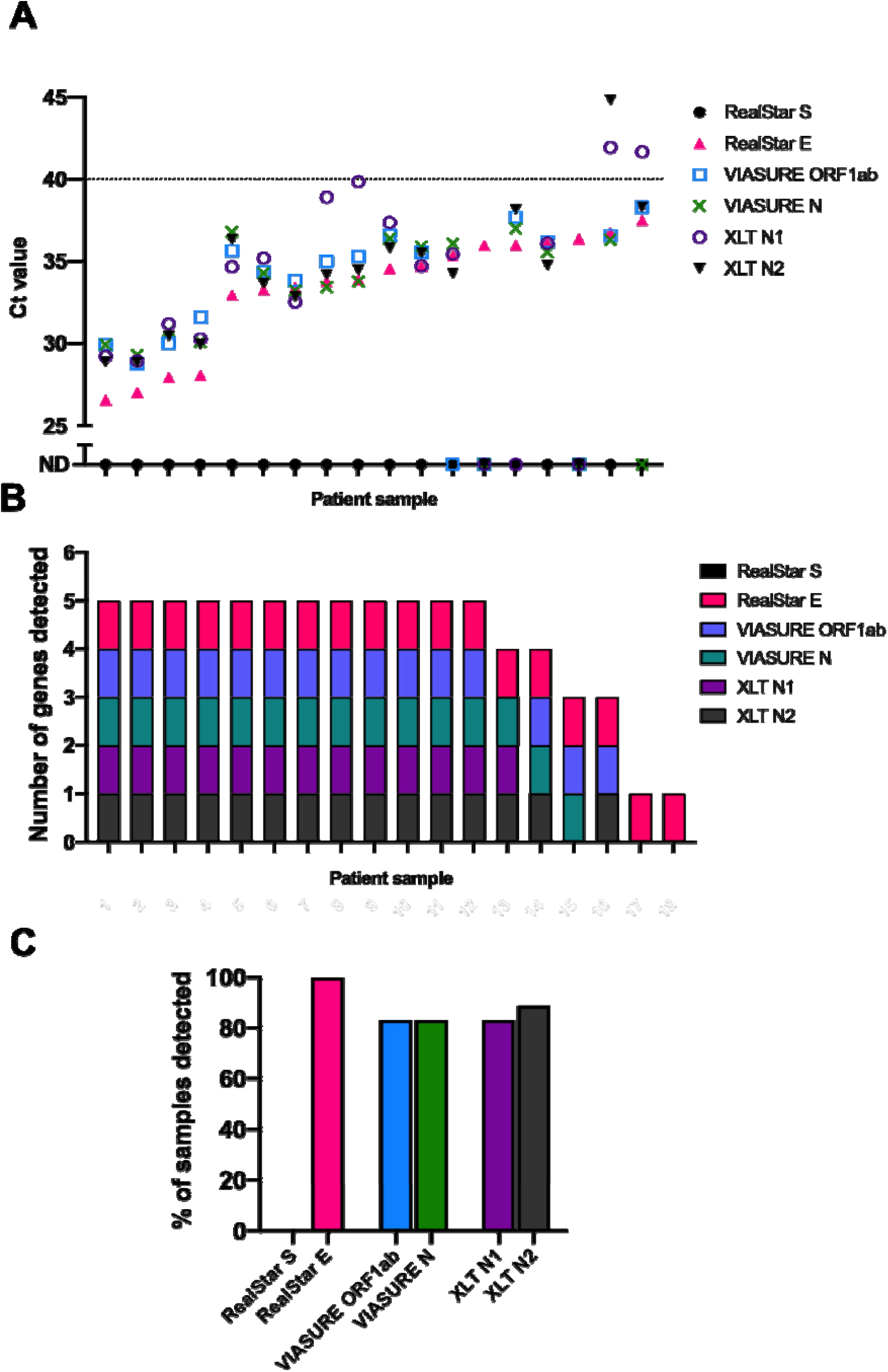
Diagnostic kit comparison on borderline SARS-CoV-2 positive RNA samples. (A) C_T_ values of equivocal samples from three diagnostic workflows; RealStar, VIASURE and Quantabio XLT. Samples with C_T_ values above the dotted line, or ND (not detected) are interpreted as negative for SARS-CoV-2 RNA (B) Total number of assay-probe sets that detected SARS-CoV-2 RNA per borderline sample. (C) Percentage of borderline samples detected with each assay-probe set. (n=18 purified patient samples).

Samples were blinded, processed and evaluated for each target. After unblinding, and as expected, the RealStar assay identified 18/18 (100% samples) positive for the E gene, but 0/18 (0%) for the S gene, corroborating these as equivocal samples provided by the clinical pathology laboratories (Table 2, Figure 2). Cross comparison between the three kits showed interesting trends. Whilst there was variability in the sensitivity of different targets between individual samples (Figure 2A), the average C_T_ values across the five positive targets was remarkably similar (range, 33.3 to 34.4; Table 2). In contrast with the RealStar system, which classed all 18 samples as equivocal (positive for E gene but not S gene), VIASURE ORF1ab and N targets were detected in 15 samples, with 14 samples being positive for both - i.e. 14 positive (77%), 2 equivocal (11%), 2 negative (11%). Similarly, the N1 and N2 targets Quantabio XLT system were positive in 15/18 and 16/18 cases respectively, with 15 samples being positive for both - i.e. 15 positive (83%), 1 equivocal (6%), 2 negative (11%). Thus, all three systems successfully identified samples from patients with a low viral load, although there were quantitative differences in the number scored as positive, equivocal or negative, showing these samples remain a diagnostic challenge. However, as samples were deemed equivocal using the RealStar assay we are aware this could demonstrate some bias in the interpretation.

### Extraction-free detection of SARS-CoV-2

A further key limitation when upscaling testing for COVID-19 (both at the local and national level) is the shortage of both commercially available viral inactivation reagents, such as GT, and RNA extraction kits. To overcome this bottleneck, several studies have shown the efficacy of heat-treating samples to 70 °C for 5 mins (6, 8) both to inactivate the virus and release encapsidated viral RNA. To investigate the impact of heat inactivation on subsequent viral detection, we obtained unprocessed aliquots from patient samples, subjected them to heat treatment and analysed using the VIASURE and Quantabio XLT methodologies. In addition, we used a second Quantabio reaction mix – UltraPlex 1-Step ToughMix, a concentrated version of the XLT mix, to minimise potential inhibitory issues with non-extracted material. We set out to determine if heat treating samples altered the outcome of the RT-qPCR results for samples previously scored as positive or negative.

The aliquots from five samples (3 positive, 2 negative) were heated to 75 °C for 10 mins and analysed using either the VIASURE, UltraPlex or XLT assays, hence avoiding chemical inactivation or dedicated RNA extraction. C_T_ values were compared against values obtained from RNA purified and analysed by the standard clinical process using the RealStar assay (Table 3).

**Table 3.** Heatmap of Ct values obtained from diagnostic kits using heat treated (75 _o_C 10 mins) swab samples (n=5 patient samples).

| Sample | RealStar<sup>a</sup> |  | Ultrplex<sup>†</sup> |  | XLT<sup>†</sup> |  | VIASURE |  |
| --- | --- | --- | --- | --- | --- | --- | --- | --- |
|  | S | E | N1 | N2 | N1 | N2 | ORF1ab | N |
| Positive 1 | 20.25 | 18.14 | 22.66 | 23.2 | 25.14 | 25.58 | 33.71 | 32.14 |
| Positive 2 | 26.14 | 23.49 | 26.63 | 27.26 | 29.64 | 30.1 | 37.21 | 35.2 |
| Positive 3 | 31.71 | 29.57 | 34.26 | 35.18 | >40 | 38.41 | >40 | >40 |
| Negative 1 | >40 | >40 | >40 | >40 | >40 | >40 | >40 | >40 |
| Negative 2 | >40 | >40 | >40 | >40 | >40 | >40 | >40 | >40 |

\* Clinical RealStar assay was performed using RNA Samples were frozen at -80 °C after heat treatment $C_T$ value > 40 =negative for SARS-CoV-2 RNA

Surprisingly, heat-inactivated samples analysed using the VIASURE assay showed C_T_ values of >10 higher (i.e. >1000-fold lower sensitivity) than values obtained from purified RNA using the RealStar assay. Moreover, the VIASURE system only detected in 2/3 (67%) of samples as positive. In contrast, the CDC N2 primer-probe coupled with the Quantabio UltraPlex and XLT one-step assays concordantly identified all positive and negative samples. However, both accuracy and sensitivity of the XLT assay were blunted relative to the UltraPlex assay. Thus, when using the XLT, the N1 probe failed to amplify one of the positive samples, whilst the UltraPlex assay gave C_T_ values of approximately 3 cycles lower (∼8-fold more sensitive) in the positive samples. Therefore, the Quantabio UltraPlex assay was used for subsequent analysis.

### Heat treatment bypasses the need for RNA extraction

To further assess the accuracy of the UltraPlex assay, in addition to the 5 samples above, we prepared aliquots of a further 6 known positive and 2 known negative samples (identified by clinical RealStar assay). Thus, the cohort was 13 samples (9 positives, 4 negatives) that had each been heated to 75 °C for 10 mins. Processing and analysis using the UltraPlex assay revealed that the N1 probe showed 100% accuracy, whereas the N2 probe detected 8/9 (89%) positive samples (Table 4). However, it should be noted, two of the samples did not amplify the RealStar S gene, suggesting that incorporating a different probe with the same UltraPlex workflow could increase accuracy.

**Table 4:** Detection sensitivity of heat-treated swab samples detected using UltraPlex assay. (n=13 patient samples)

| Probe | Results (n) |  | Accuracy (%) |
| --- | --- | --- | --- |
|  | TP | TN |  |
| N1 | 9/9 | 4/4 | 100.0 |
| N2 | 8/9 | 4/4 | 88.9 |

TP, true positive; TN, true negative

We then carried out a direct comparison of heat treatment versus traditional RNA purification protocols using the UltraPlex assay. We analysed purified RNA from 7 of the above positive and 2 negative samples with the UltraPlex assay. The majority of samples showed similar C_T_ values after heat treatment compared to RNA extraction; however one sample demonstrated early amplification for both N1 and N2 genes when heat-treated implying better detection of virus without RNA extraction. All heat-treated and RNA extracted samples were in concordance for the N2 gene. Surprisingly, N1 was amplified in every heat-treated sample, whereas one RNA extracted sample failed to be detected by the same probe (Figure 3). Nevertheless, the results in Table 5 suggest that no single assay is superior in all instances and as the C_T_ values increase there is more variation between assays and consequently the choice of assay may become more important when detecting samples with low viral load.

**Table 5:** Heatmap to show direct comparison of heat-treated and RNA extracted Ct values using UltraPlex and clinical RealStar assays (n=7).

| Sample | Ultrplex N1 |  | Ultrplex N2 |  | RealStar® |  |
| --- | --- | --- | --- | --- | --- | --- |
|  | RNA | HI | RNA | HI | S | E |
| 1 | 21.21 | 21.23 | 21.38 | 21.93 | 19.05 | 17.11 |
| 2 | 22.31 | 20.04 | 24.22 | 21.07 | 22.49 | 19.31 |
| 3 | 36.52 | 26.63 | 34.24 | 27.26 | 26.14 | 23.49 |
| 4 | >40 | 26.51 | 27.81 | 27.09 | 26.42 | 24.41 |
| 5 | 31.08 | 32.09 | 31.06 | 32.79 | 29.33 | 26.88 |
| 6 | 36.55 | 37.98 | 36.36 | 38.24 | >40 | 33.23 |
| 7 | 38.85 | 38.9 | >40 | >40 | >40 | 38.4 |

\* RealStar assay was performed using RNA C<sub>T</sub> value > 40 =negative for SARS-CoV-2 RNA

**Figure 3:**
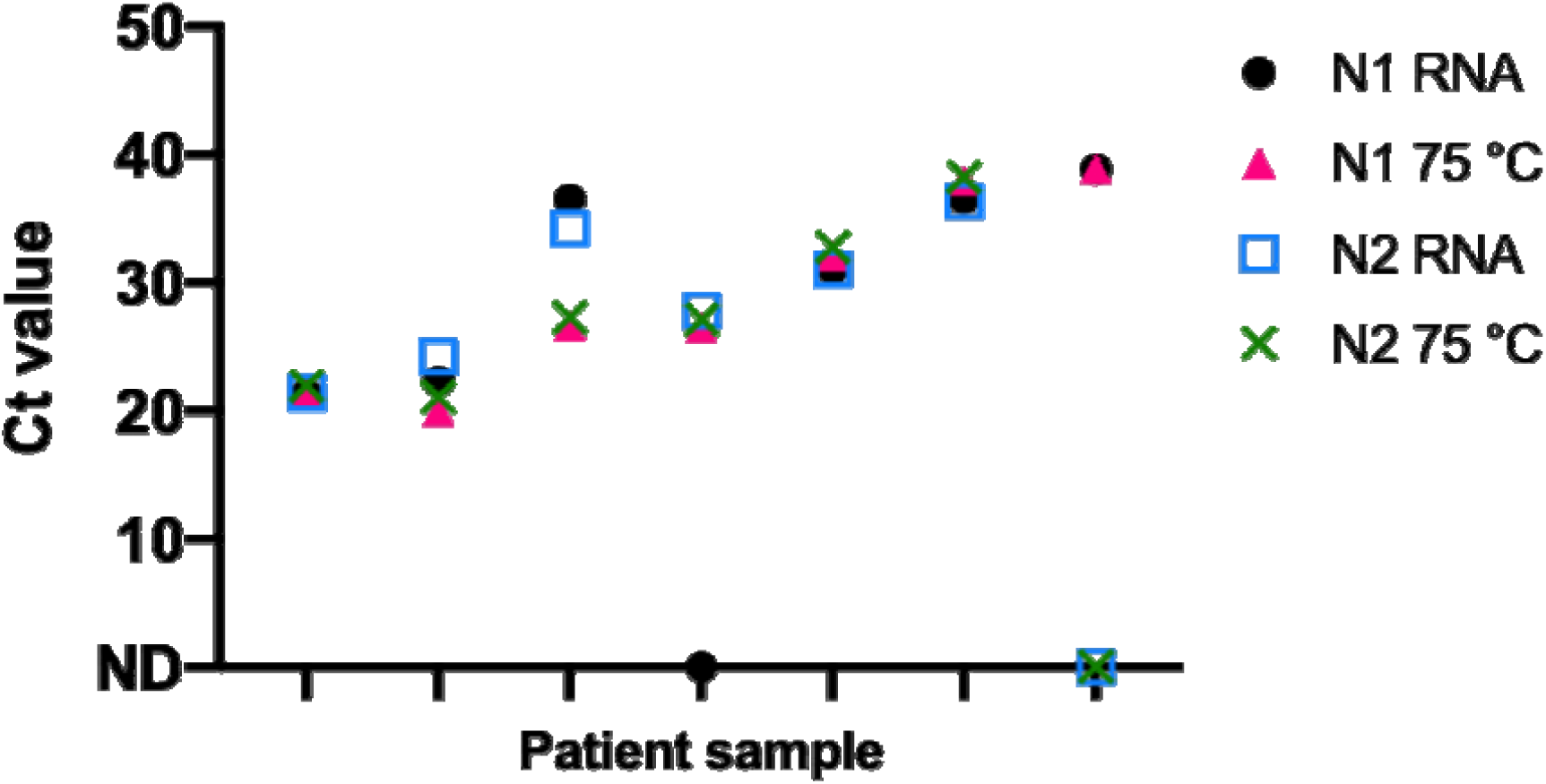
Ct value comparison of heat inactivated material and extracted RNA from the same swab samples. Samples were heat treated at 75 °C for 10 minutes. RT-qPCR run with two primer-probe sets (N1 and N2) using Quantabio UltraPlex assay (n=7 patient samples).

Having shown heat treatment was comparable to RNA purification, we wanted to test the UltraPlex assay on a larger set of samples. 129 samples were provided by PLP. These had been heat treated at 90-95 C for 10 min, with subsequent RNA extraction prior to processing via the Abbott system. This method of choice at PLP determined 109 of these samples as positive and 20 as negative. We repeated testing of these samples using the UltraPlex assay on the heat-inactivated material. Two samples were inconclusive (the internal cellular RNaseP gene did not amplify) and so were removed from analysis. The remaining samples gave an accuracy of 88.98% and 92.91% for the N1 and N2 probes respectively (Table 6). Importantly, whilst 2 false positives were detected with the N1 probe, no false positives were detected with the N2 probe. However, both probes showed some false negatives.

**Table 6:** Detection performance of N1 and N2 UltraPlex assay-probe sets on heat-treated swab samples, compared to Abbott assay results from Lincoln.

| Probe | Results (n) | | | | Accuracy (%) | Avg Ct shift ± SEM |
| --- | --- | --- | --- | --- | --- | --- |
|  | TP | TN | FP | FN |  |  |
| N1 | 95 | 18 | 2 | 12 | 88.98 | 12.54 ± 0.29 |
| N2 | 98 | 20 | 0 | 9 | 92.91 | 13.52 ± 0.18 |

TP, true positive; TN, true negative; FP, false positive; FN, false negative.

When looking at the raw C_T_ values, we noted a rightward shift of 12.5 cycles for the Ultraplex N1 probe and 13.5 for the N2 probe relative to the data provided by PLP using the Abbot system (Table 6; Figure 5). This is to be expected since signal intensity on the Abbot system is very high relative to other commercial kits on account of dual viral target identification (RdRp and N-genes). Notwithstanding this technical difference, there was a strong positive correlation between the two systems (N1 probe R_2_=0.8020; N2 probe R_2_=0.9259; Figure 4). Altogether, these data confirm that the UltraPlex system provides an accurate and sensitive testing approach for heat-extracted RNA samples, with data comparing favourably in comparison with other platforms and extraction methods.

**Figure 4:**
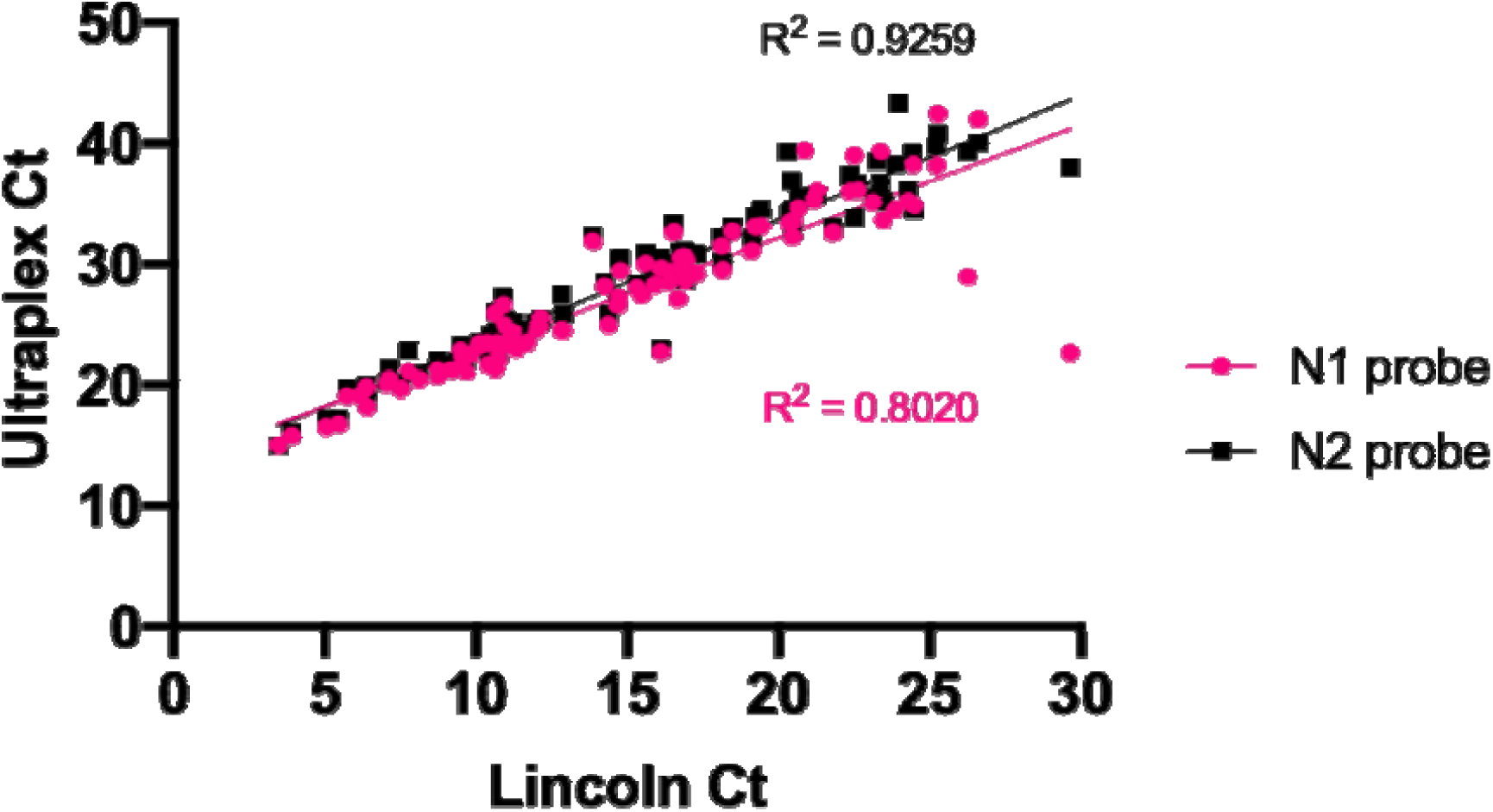
Correlation between CT values obtained by N1 and N2 UltraPlex assay-probe sets and Abbott assay (n=127). Linear regression and R^2^ correlation values for each probe are indicated

## Discussion

Efficient diagnostic testing for SARS-CoV-2 virus has been an essential element in tackling the COVID-19 pandemic and will continue to be so with ‘test, track and trace’ strategies operational in many countries. The rapid, global spread of the virus was met with a necessity for large-scale SARS-CoV-2 testing. Very quickly, this revealed vulnerabilities in production and distribution lines for RNA extraction reagents, assays, and equipment. To provide contingency for increased local diagnostic capacity and circumvent emerging bottlenecks in testing consumables supply chains, we assessed two commercially available diagnostic workflows (CerTest VIASURE kit and Quantabio one-step RT-qPCR assays combined with CDC primer-probe sets). Both assays showed concordance in diagnostic accuracy with the routinely used and validated Altona RealStar® assay for COVID-19 diagnosis in NUH clinical laboratories; this is similar to previous published results (9). Whilst full agreement in assigning positivity and negativity was observed for the true positive and negative samples, equivocal samples as defined by the clinical RealStar® assay, generated differing results in alternative assays. The different genes targeted by the individual assays is likely to rely on the differential expression of the genes (10, 11). The variability suggests that no one system will preferentially detect all weakly positive samples, but rather the detection ability will be sample and replicate dependent.

Alternative testing methods to the widely used two-step RNA purification and RT-qPCR, are being employed by laboratories to increase capacity and avoid limitations caused by supply constraints (9). We confirm viral inactivation using heat treatment and direct RT-qPCR, bypassing RNA extraction, can be used to detect SARS-CoV-2 in swab samples. However, we found accurate detection using this method to be dependent on the RT-qPCR assay used. We thus highlight the need for local assay validation before implementing an extraction-free workflow in a clinical laboratory.

The VIASURE assay performed well on purified RNA, but was inhibited by non-purified material. Further investigation into the incompatibility of the VIASURE assay with crude swab material, using spiked RNA, revealed the reverse transcriptase step is adversely affected (data not shown). This could be explained by autofluorescence of the viral transport medium, or other contaminants within swab samples. In contrast to the VIASURE assay, the combined CDC N1, N2 Quantabio UltraPlex assay gave comparable results using both RNA purification and extraction-free methods. Similar discrepancies between assay kits have been reported in other heat-processing systems (12). Detection accuracy of viral RNA swabs in clinical laboratories varies on the swab type used, the quality of sample collection and the stage of the viral infection. The combined CDC N1, N2 Quantabio UltraPlex assay was able to detect SARS-CoV-2 in a large cohort of swab samples that had been collected from various testing sites. Furthermore, these samples had undergone different heat treatments (75 °C or 90-95 °C for 10 mins), showing flexibility in application of the combined CDC N1,N2 UltraPlex assay in laboratories which employ either temperature during sample processing. During the writing of this paper, CDC evaluated the Quantabio UltraPlex kit with heat treatment and it has been granted FDA approval for use in some scenarios (www.fda.gov/media/134922/download).

A limitation of the commercially available CDC probes is use of the same reporter for all targets, thus requiring three separate reactions per patient sample (for each N1, N2 and RNaseP target). Perchetti et al recently reported the ability to multiplex these targets, with no loss in detection sensitivity (13). We have taken a similar approach and, moving forwards, have chosen to triplex the N2, E and RNaseP targets, to increase testing throughput. We have chosen the N2 target as this was more sensitive than the N1 target when detecting SARS-CoV-2 RNA in heat treated swab samples. In this study, there were a significant number of samples analysed by the RealStar assay that only had amplification of the E gene, and not the S gene. Thus, the E target has been included in order to detect additional positives. Furthermore, multiplexing of N2 and E targets without loss of sensitivity has previously been reported (14).

A recent study suggests testing frequency matters more than sensitivity, particularly in large communities e.g. universities where COVID-19 could spread rapidly (15). However, it might be more important to favour sensitivity and specificity over regular testing in order to reduce the risk of false negative results, especially when testing an asymptomatic population. Virus transmission also occurs in asymptomatic individuals, which further necessitates the ability to perform large-scale and rapid testing (16, 17). Bypassing the requirement for RNA extraction significantly decreases both cost and time required for assay completion. It also makes testing more amenable to developing areas without the infrastructure required for high throughput RNA extraction. We have demonstrated the feasibility of using heat treatment with a one-step RT-qPCR assay to reliably diagnose COVID-19.

## Data Availability

NA

## Acknowledgement

The University of Nottingham qPCR team which to thank Dr Hiten Mistry, Maria Barbadillo-Munoz and Carl Winfield for their efforts in making our laboratory safe, and the resources available during the lockdown time. We also thank the supervisors and Principal Investigators of the lead authors for their support in allowing their staff to contribute to this project. Also the laboratory staff working on SARS-C0V-2 testing at Nottingham University Hospitals NHS Trust for processing the many samples included in this study. Funding for this project was from the University of Nottingham. JLT is funded by EPSRC grant EP/N006615/1. AVB is funded by the British Heart Foundation (PG/18/31/33759) and Royal Society (RGS\R1\191221) CS receives funding from Children’s Cancer and Leukaemia Group – Little Princess Trust (CCLGA 2019 2) and Cancer Research UK (C11392/A27985)

## Notes

### Competing Interest Statement

The authors have declared no competing interest.

### Funding Statement

EPSRC grant EP/N006615/1

British Heart Foundation (PG/18/31/33759)

Royal Society (RGS\R1\191221) Childrens Cancer and Leukaemia Group Little Princess Trust (CCLGA 2019 2)

Cancer Research UK (C11392/A27985)

